## Supplementary text for "Cost-effectiveness of Dengue Vaccination in Puerto Rico"

### SUPPORTING INFORMATION

#### Sensitivity analysis

##### *Higher transmission setting*

We explored a scenario in which Puerto Rico has a higher transmission intensity of $PE_9 = 60\%$. In this scenario, we found that around five hospitalizations in naïve children occurred for every 1,000 people vaccinated, compared to six hospitalizations with $PE_9 = 50\%$ (Figure S1). Compared to the baseline scenario, we found a slight increase in the number of hospitalizations averted for each additional hospitalization in the naïve vaccinated group. However, in the scenario of higher specificity (0.99) with lower sensitivity (0.64), we found that this proportion of hospitalizations averted almost tripled, resulting in 141 (84 - 813) hospitalizations averted for each additional hospitalization in the naïve vaccinees group.

We found that the intervention was slightly more cost-effective in a higher transmission setting of $PE_9 = 60\%$ (Figure S2). The ICER of the intervention was around 90,000 USD per QALY averted, which represented a reduction of 18,000 USD from the baseline scenario of $PE_9 = 50\%$. In terms of symptomatic cases and hospitalizations, the ICER was also lower. We estimated an ICER of 8,000 USD per symptomatic case averted, and 11,000 USD per hospitalization averted.

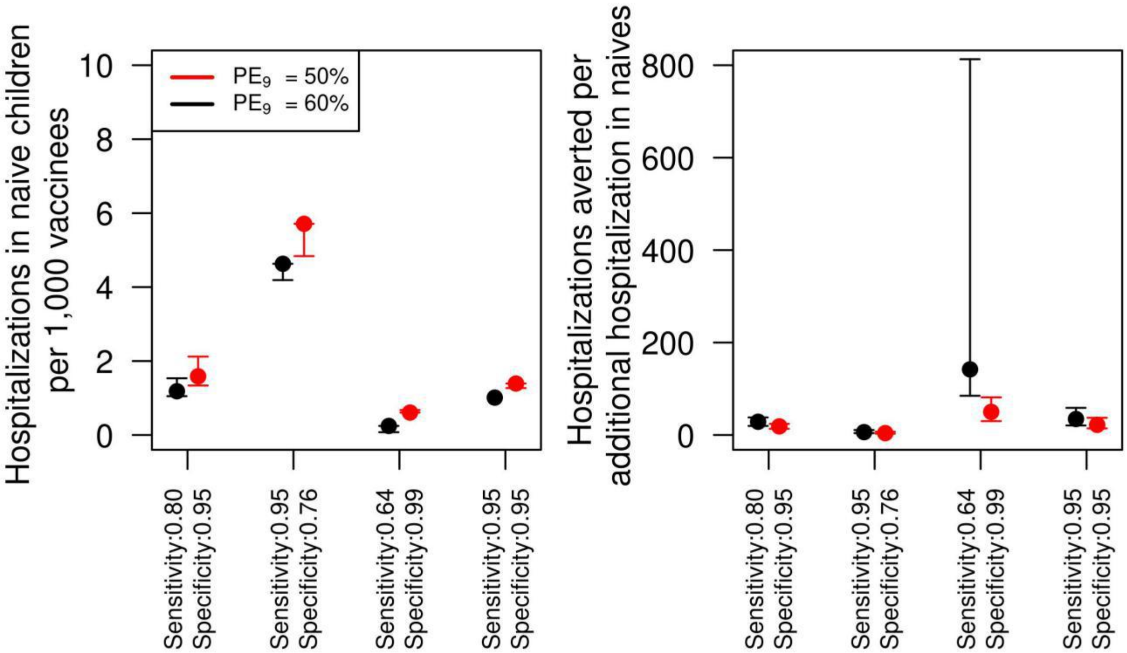


Figure S1. Number of additional hospitalization cases due to vaccination of DENV-naïve children and ratio of hospitalizations averted to additional hospitalizations at different levels of sensitivity and specificity. Left panel shows the number of hospitalizations per every 1,000 children vaccinated. The right panel shows the number of hospitalizations averted for every additional hospitalization case in the DENV-naïve group. The simulations were performed for 80% intervention coverage of routine pre-vaccination screening in 9 year-olds over 10 years.

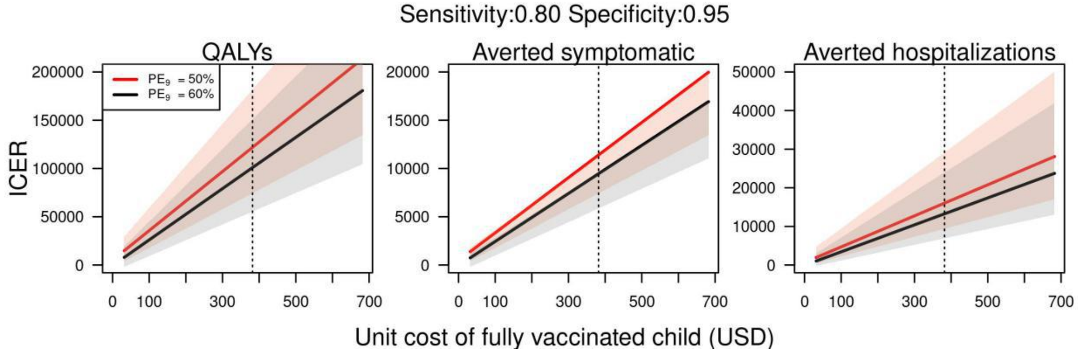


Figure S2. ICER of pre-vaccination screening strategy in Puerto Rico with a higher transmission setting ($PE_9 = 60\%$) at different costs of vaccination (total cost for three doses per person), assuming a unit cost of serological screening of 30 USD. Red lines represent a transmission intensity scenario of $PE_9 = 50\%$, and black lines represent a transmission scenario of $PE_9 = 30\%$. All costs in 2019 USD.

***Sensitivity of cost-effectiveness to uncertainty in sensitivity and specificity values***

The cost-effectiveness ratio of the intervention increased in a scenario of lower specificity (0.76) with higher sensitivity (0.95). The increase was higher in a low-transmission scenario, in part because a reduction in specificity in such a low-transmission level implied a lower number of hospitalizations averted and a higher proportion of hospitalizations caused by misclassification. In contrast, increasing the specificity while reducing sensitivity, slightly reduced the ICER. Changes in the sensitivity and specificity of serological screening did not affect substantially the cost to avert a symptomatic case. With an assumption of lower specificity, more hospitalized cases occurred in the lower transmission scenario, increasing the cost to avert a hospitalization to around 66,000 USD. The cost to avert a hospitalization also increased in a moderate transmission setting to around 24,000 USD. Finally, increasing specificity at a lower sensitivity reduced the cost per hospitalization averted to around 29,000 USD for a low transmission setting.

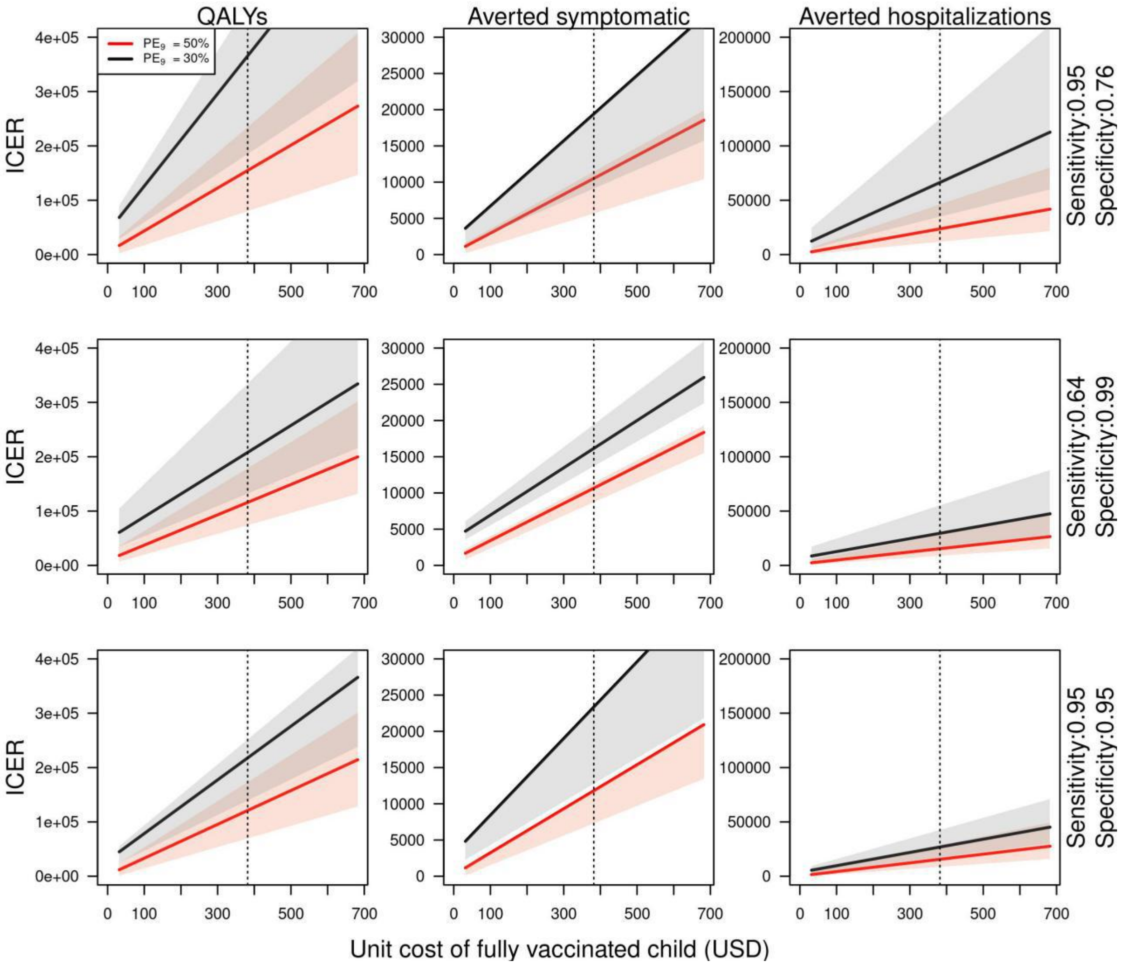


Figure S3. ICER of pre-vaccination screening strategy in Puerto Rico at different cost of vaccination (3 doses per person), assuming a unit cost of serological screening of 30 USD. Dotted vertical line represents the baseline cost of vaccination (382 USD). All costs in 2019 USD.

***Lower coverage***

Achieving 80% coverage of the intervention (i.e., serological screening and vaccination in the event of a positive result) in 9-year-olds might be unfeasible. We explored a scenario with a lower coverage of 50% to estimate the effects on cost-effectiveness at a lower vaccination coverage. We found that a lower coverage of vaccination increases slightly the incremental cost of gaining a QALY in a moderate transmission setting ($PE_9 = 50\%$)(Fig. S4, left panel). Lower coverage also slightly increased the cost to avert a symptomatic case. In contrast, lower coverage had a minimal impact on the cost-effectiveness to avert a hospitalization case (Fig. S4). Although we assume that the vaccine does not provide permanent protection against infection, the slight difference in the cost-effectiveness at lower coverage could be explained by the temporary cross-protection acquired from vaccination, resulting in indirect protection from vaccination in the short term. The overall magnitude of this indirect protection increases with coverage, as more people acquire temporary protection from infection. Given that some cases would be prevented from this indirect protection, in a moderate transmission setting the high-coverage scenario has slightly lower cost per averted symptomatic case than the low-coverage scenario.

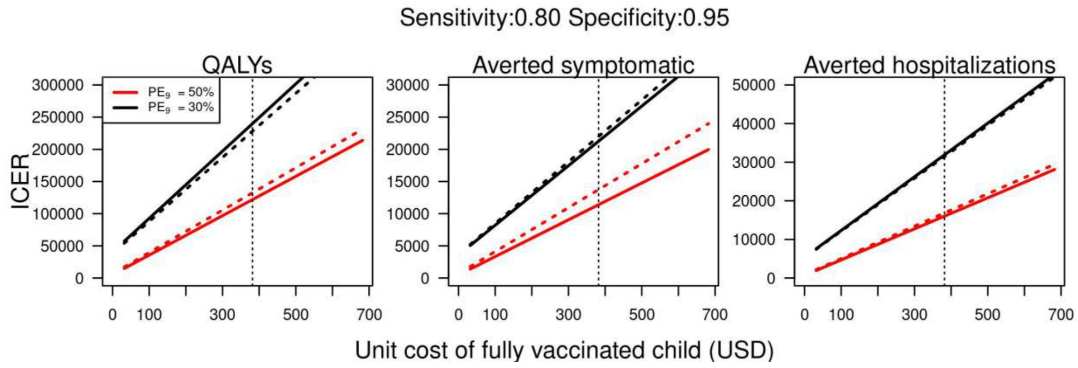


Figure S4. ICER of pre-vaccination screening strategy in Puerto Rico at lower coverage (50%) and at different cost of vaccination (3 doses per person), assuming a unit cost of serological screening of 30 USD. Solid lines shows the baseline scenario of coverage (80%) and dashed line shows lower coverage assumption (50%). All costs in 2019 USD.

***Uncertainty about costs and disutility values***

To reflect uncertainty on the assumptions of treatment costs and disutility, we varied the costs of hospitalized and symptomatic cases by 20%, and used the uncertainty intervals on disutility values from Zeng et al. [26] (Table S1). Reducing the costs of hospitalization 20% below the baseline assumption resulted in an increase of around 125,000 USD per QALY gained in the ICER, while an increment of 20% reduced the ICER to 119,000 USD. Estimates of the ICER showed little sensitivity to 20% variation of the cost of clinical attention of symptomatic cases (121,000 USD - 122,500 USD). Uncertainty on the disutility values of symptomatic cases resulted in a difference of around 62,000 USD in the ICER, while the uncertainty on the disutility of hospitalizations resulted in a 44,000 USD difference in the ICER. Similar magnitudes were found for the sensitivity of the ICER estimates at lower transmission intensity. Given the large uncertainty associated with the cost of serological screening, we assumed a wide range of values with a lower bound of the unit cost of serological screening of 1 USD, and upper bound of 60 USD. The ICER on these upper and lower bounds showed that increasing the cost to 60 USD increased the ICER to 143,000 USD, whereas the ICER would be reduced to 101,000 USD with an assumption of 1 USD.

Table S1. Sensitivity to changes in costs and disutility for ICERs measuring costs (2019 USD) per QALYs gained.

| Parameter | $PE_9$ | Min | ICER(Min) | Max | ICER(Max) |
|---|---|---|---|---|---|
| costs symptomatic | 50% | 252 | 122,533 | 379 | 121,358 |
| costs hospitalization | 50% | 1,706 | 124,664 | 2,559 | 119,228 |
| unit cost of serological screening | 50% | 1 | 101,068 | 60 | 143,541 |
| disutility symptomatic | 50% | 0.0170 | 139,192 | 0.0917 | 78,538 |
| disutility hospitalization | 50% | 0.0241 | 131,241 | 0.0960 | 87,609 |
| costs symptomatic | 30% | 252 | 240,597 | 379 | 239,433 |
| costs hospitalization | 30% | 1,706 | 242,749 | 2,559 | 237,281 |
| unit cost of serological screening | 30% | 1 | 184,912 | 60 | 297,008 |
| disutility symptomatic | 30% | 0.0170 | 274,684 | 0.0917 | 151,714 |
| disutility hospitalization | 30% | 0.0241 | 257,317 | 0.0960 | 172,741 |

**Vaccine cost estimates**

Table S2. Estimated price for CYD-TDV based on age range, recombinant technology, and recent approval.

| | Cost/dose | Cost/ 3 doses |
|---|---|---|
| Estimated cost per dose based on three factors (age range, recombinant technology, recent approval): | 107.28 | 321.8 |
| Lowest observed: | 10.85 | 32.6 |
| Highest observed: | 227.93 | 683.8 |
| Average private sector cost per dose of vaccines recommended for 7 to 15 year olds: | 113.54 | |
| Average private sector cost per dose of recombinant vaccines: | 85.13 | |
| Average private sector cost of vaccines approved by FDA since 2014: | 123.16 | |

Table S2. List of vaccines recommended for 7 to 15 year-olds[1].

| Vaccines recommended for 7 to 15 year olds (i.e., close in age to 9 year olds) | Private sector cost/dose |
|---|---|
| Meningococcal ACWY-D, ACWY-CRM | |
| Menveo | 130.75 |
| Menactra | 122.31 |
| Tetanus, diphtheria, & acellular pertussis | |
| Boostrix | 41.19 |
| Adacel | 45.5 |
| Human papillomavirus | |
| Gardasil9 | 227.931 |
| **Average** | **113.54** |

[1] Source: All Private sector cost/dose from the Vaccine Price List

(https://www.cdc.gov/vaccines/programs/vfc/awardees/vaccine-management/price-list/index.html) last updated on 12/2/2019

Table S3. List of vaccines with similar technology as CYD-TDV

| Vaccines that are subunit/recombinant technology (i.e., similar technology as Dengvaxia) | Private sector cost/dose |
|---|---:|
| Hib (Haemophilus influenzae type b) | |
| PedvaxHIB | 26.333 |
| ActHIB | 16.51 |
| Hiberix | 10.85 |
| Hepatitis B | |
| Engerix B | 23.72 |
| Recombivax HB | 23.95 |
| Human papillomavirus | |
| Gardasil9 | 227.931 |
| Pertussis | |
| Boostrix | 41.19 |
| Adacel | 45.5 |
| Pneumococcal | |
| Prevnar 13 | 188.26 |
| Pneumovax | 105.194 |
| Meningococcal | |
| Menveo | 130.75 |
| Menactra | 122.31 |
| Shingles | |
| Shingrix | 144.2 |
| **Average** | **85.13** |

Table S4. List of vaccines introduced in the last 5 years[2].

| Vaccines introduced in the last 5 years (i.e., similar time-frame) | private sector cost/dose | FDA approval |
|---|---|---|
| Shingrix | 144.2 | 2017 |
| Heplisav-B | 115.75 | 2017 |
| Hiberix | 10.85 | 2016 |
| Vaxchora | | 2016 |
| Bexsero | 170.75 | 2015 |
| Quadracel | 53.13 | 2015 |
| Fluad | | 2015 |
| Trumenba in the U.S. to prevent serogroup B meningococcal disease | 139.52 | 2014 |
| Gardasil 9 (Merck) | 227.931 | 2014 |
| **Average** | **123.16** | |

[2] Sources: All cost per dose information comes from the VFC vaccine price list (https://www.cdc.gov/vaccines/programs/vfc/awardees/vaccine-management/price-list/index.html). All FDA approval information comes from immunize.org timeline (https://www.immunize.org/timeline/)
